# Polygenic Risk Score Stratification Guides Prioritization of Modifiable Risk Factors for Stroke Prevention

**DOI:** 10.64898/2026.06.28.26356786

**Authors:** Yoichi Sutoh, Tsuyoshi Hachiya, Yayoi Otsuka-Yamasaki, Motoki Nakao, Shinichi Omama, Yorihiko Koeda, Hiroshi Akasaka, Yuka Kotozaki, Shohei Komaki, Shiori Minabe, Hideki Ohmomo, Naoyuki Nishiya, Makoto Sasaki, Kozo Tanno, Atsushi Shimizu

**Affiliations:** Iwate Tohoku Medical Megabank Organization, Iwate Medical University, Yahaba, Japan; Institute for Biomedical Sciences, Iwate Medical University, Yahaba, Japan; Tohoku Medical Megabank Organization, Tohoku University, Sendai, Japan; Department of Pharmacology, School of Medicine, Iwate Medical University, Sapporo, Japan; Department of Cardiovascular Medicine, Faculty of Medicine and Graduate School of Medicine, Hokkaido University, Sapporo, Japan; Department of General Medicine, School of Medicine, Iwate Medical University, Yahaba, Japan; Division of Cardiology, Department of Internal Medicine, Iwate Medical University, Yahaba, Japan; Department of Hygiene and Preventive Medicine, School of Medicine, Iwate Medical University, Yahaba, Japan; Department of Clinical Pharmacy, Division of Integrated Information for Pharmaceutical Sciences, School of Pharmacy, Iwate Medical University, Yahaba, Japan

**Author notes:** Corresponding authors: Yoichi Sutoh, PhD Division of Biobank Utilization and Facilitation, Iwate Tohoku Medical Megabank Organization, Iwate Medical University 1-1-1 Idaidori, Yahaba, Shiwa, Iwate 028-3694, Japan, ex. 5471, Tsuyoshi Hachiya, PhD Iwate Tohoku Medical Megabank Organization, Iwate Medical University 1-1-1 Idaidori, Yahaba, Shiwa, Iwate 028-3694, Japan, Atsushi Shimizu, PhD Tohoku Medical Megabank Organization, Tohoku University 2-1, Seiryo-machi, Aoba-ku, Sendai 980-0872, Japan, ex. 3684. Equally contributed (Yoichi Sutoh, Tsuyoshi Hachiya).

**Keywords:** PGS, Stroke, lifestyle, modifiable risk, prospective study

## Abstract

**Background:** Integrating polygenic scores (PGS) into population-based stroke prevention strategies may help reduce the stroke burden at both individual and population levels. However, prospective evidence regarding the interaction between genetic and modifiable risk factors remains limited, particularly in East Asian populations.

**Methods:** We evaluated the influence of modifiable risk factors across PGS levels in the prospective population-based Tohoku Medical Megabank Community-Based Cohort Study. Stroke PGS was calculated using a model developed by the GIGASTROKE project.

**Results:** During a median follow-up of 4.84 years (121,843.2 person-years), 246 incident stroke events were identified. Compared with participants in the intermediate PGS group (third quintile), those in the highest PGS group (fifth quintile) had a 60% greater risk of incident stroke (hazard ratio [HR], 1.60; 95% confidence interval [CI], 1.10–2.34). When participants with intermediate genetic risk and no modifiable risk factors were used as the reference group, hypertension (HR, 2.17; 95% CI, 1.11–4.25) and diabetes (HR, 2.10; 95% CI, 1.04–4.27) were significantly associated with an increased stroke risk in the intermediate PGS group. In contrast, multiple modifiable risk factors were significantly associated with stroke risk in the highest PGS group, including hypertension (HR, 2.90; 95% CI, 1.53–5.48), diabetes (HR, 2.07; 95% CI, 1.02–4.20), dyslipidemia (HR, 2.20; 95% CI, 1.28–3.79), obesity (HR, 1.98; 95% CI, 1.19–3.30), and smoking (HR, 2.57; 95% CI, 1.34–4.89). Participants with both the highest PGS and ≥ 4 modifiable risk factors had a substantially elevated stroke risk (HR, 2.76; 95% CI, 1.28–5.92). The combination of genetic and modifiable risk factors significantly influenced the cumulative incidence of stroke (*P*<0.001).

**Conclusions:** Modifiable risk factors substantially influenced stroke risk even among participants with high genetic susceptibility. The relative importance of modifiable risk factors for stroke prevention may differ according to an individual’s genetic risk level.

## Introduction

Stroke remains a major cause of mortality and the third leading cause of disability worldwide^1–4^. Although age-standardized incidence rates of stroke have substantially declined over the past three decades, the absolute numbers of stroke cases and stroke-related deaths have steadily increased, particularly in aging populations^3,4^. Given the substantial heritability of stroke^5–10^, identifying individuals at high genetic risk and implementing targeted lifestyle interventions may represent a promising strategy for primary prevention of stroke^11–14^.

The GIGASTROKE project, the largest genome-wide association study (GWAS) of stroke, conducted a cross-ancestry meta-analysis involving approximately 110,000 cases and 1.5 million controls, which identified 89 susceptibility loci^15^. Based on these findings, polygenic scores (PGS) for stroke have been developed and validated in European and East Asian populations. The predictive performance of the PGS in East Asian populations was comparable to that observed in European populations. For example, individuals in the highest PGS quintile had a 1.64-fold greater risk of ischemic stroke than those in the middle quintile, whereas those in the top 5% had a 1.94-fold greater risk. Despite ongoing challenges in trans-ancestry portability and predictive accuracy, the clinical application of stroke PGS is becoming increasingly feasible.

Several studies have investigated the relationship between genetic risk and lifestyle factors^16–18^. However, prospective evidence remains limited, particularly in East Asian populations. Therefore, further evidence is needed to guide the integration of PGS into population-based prevention strategies^19^.

In this study, using data from one of the largest prospective community-based cohorts in Japan, we assessed the potential of modifiable risk factors to mitigate stroke risk among individuals with a high genetic predisposition to stroke.

## Methods

### Study population

We conducted a prospective 5-year follow-up study of participants enrolled in the Tohoku Medical Megabank Project community-based cohort (TMM CommCohort). Details of this cohort have been described previously^20–22^. Briefly, the cohort comprised more than 87,000 adults aged 20–75 years residing in northeastern Japan between May 2013 and March 2016. At baseline, participants completed questionnaires on sociodemographic characteristics, lifestyle habits, and medical history and underwent physiological examinations and blood and urine tests. Among TMM CommCohort participants, those residing in Iwate Prefecture at baseline were covered by the Iwate Prefecture Stroke Registration program^23,24^ and were therefore eligible for the follow-up of incident stroke events.

The primary endpoint of this study was the first occurrence of stroke during the follow-up period from 2013 to 2019 (median follow-up duration: 4.84 years).

Incident stroke cases were identified using the Iwate Stroke Registry, a population-based stroke registration program coordinated by the Iwate Prefectural Government and Iwate Medical Association, which covered the entire study area where the cohort participants resided^25–27^. Information on stroke events was supplemented through active surveillance survey conducted by survey teams composed of trained research nurses, research doctors, neurologists, and neurosurgeons. The medical records of hospitalized patients and deceased outpatients with cerebrovascular disease were retrospectively reviewed for inclusion in the stroke registry^25^.

Participants who did not experience a stroke event during the follow-up period and those who moved out of the study area were administratively censored at the end of follow-up or upon relocation, respectively. Investigators confirmed information on death and the date of death by reviewing the population registry records of the cohort participants^26^.

This study was approved by the Institutional Review Board of Iwate Medical University (Approval Nos. HG H25-2, HG2018-004, HG2021-011, and 2014-0001). Written informed consent was obtained from all the participants. All study procedures were conducted in accordance with the principles of the Declaration of Helsinki. This manuscript was prepared in accordance with the STROBE reporting guidelines.

### Genotyping, quality control, and genotype imputation

Genotyping, quality control, and genotype imputation have been described elsewhere^28^. Briefly, participants were genotyped using the Axiom Japonica array version 2 (JPAv2)^29^. Autosomal variants were used for quality control because the PGS models used in the present study were constructed using these variants^15^.

Quality control and genotype imputation were performed for each batch. Variants with low call rates (<0.99), Hardy–Weinberg equilibrium exact test *P*-values (<0.00001), and minor allele frequencies (<0.01) were excluded^30,31^, followed by pre-phasing and genotype imputation using SHAPEIT2 (version r837)^32^ and IMPUTE4 (version r300.3)^33^, respectively. Participants with a low call rate (<0.95) and discrepancies between self-reported and genetically inferred sex were excluded.

Principal components (PCs) for the JPAv2 genotype data were estimated using PLINK2 after standard sample- and variant-level quality control and linkage disequilibrium pruning, as described previously^34^.

### Polygenic score

We used the PGS for East Asian populations developed by the GIGASTROKE project^15^, which included 6,010,730 variants. The GIGASTROKE PGS model for East Asians was derived from GWASs of stroke and stroke subtypes, including any stroke, any ischemic stroke, large artery stroke, small vessel stroke, and cardioembolic stroke, as well as GWASs of stroke-related risk factors, including atrial fibrillation, coronary artery disease, diabetes mellitus, systolic blood pressure (SBP), diastolic blood pressure (DBP), serum total cholesterol (TC), low-density lipoprotein cholesterol (LDL-C), high-density lipoprotein cholesterol (HDL-C), triglycerides (TG), body mass index (BMI), height, and smoking behavior. The East Asian PGS model was obtained from the PGS Catalog database^35^ (PGS ID: PGS002725).

The PGS was calculated from imputed genotype dosage data and variant weights using the --score function in PLINK 2 (version 2.00a2LM)^36^ based on the following equation:

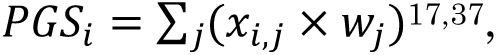

where *_PGSi_* represents the PGS for individual *i*, *_wj_* denotes the weight assigned to variant *j* (1 ≤ j ≤ 6,010,730), and *_x_*_!,#_ represents the imputed genotype dosage of variant *j* for individual *i*.

### Modifiable risk factors

Modifiable risk factors were assessed using questionnaire data and clinical examinations conducted at baseline. According to the guidelines of the Ministry of Health, Labour and Welfare of Japan, blood pressure was measured twice by trained personnel using automated devices, and the mean of the two measurements was used in the present analysis^21,31,38^. Hypertension was defined as an SBP ≥140 mmHg, a DBP ≥90 mmHg, or current use of antihypertensive medication.

Glycated hemoglobin (HbA1c) levels were measured using high-performance liquid chromatography^21,30^. Diabetes mellitus was defined as HbA1c ≥6.5% or current use of antidiabetic medications. Serum TC and TG levels were measured enzymatically^21^. Serum LDL-C and HDL-C levels were measured using the direct method. Dyslipidemia was defined as LDL-C ≥3.62 mmol/L, HDL-C <1.03 mmol/L, TG ≥1.69 mmol/L, or current use of cholesterol-lowering medication. Obesity was defined as BMI ≥25 kg/m^2^ based on measured height and weight. Current smoking and alcohol consumption were classified as either present or absent based on self-administered questionnaires.

## Statistical analysis

Participants with available genotype PC data, follow-up data (excluding those with a history of stroke at baseline), PGS data, and baseline data (including age and sex) were included in this study. Accordingly, 24,907 participants were eligible for analysis. Participants with missing data for any modifiable risk factor were excluded from the analysis.

Participants were categorized according to PGS quintiles. Trends in the means and frequencies of potential risk factors across PGS quintiles were assessed using linear and logistic regression models, respectively. Hazard ratios (HRs) and corresponding 95% confidence intervals (CIs) for incident stroke were estimated using Cox proportional hazards models adjusted for age and sex. Statistical interactions were evaluated by including multiple interaction terms in the Cox proportional hazards model. All statistical tests were two-sided, and P-values <0.05 were considered statistically significant.

All statistical analyses were performed using R version 4.5.0 (R Foundation for Statistical Computing, Vienna, Austria). To account for competing risks, cumulative incidence functions, Fine–Gray subdistribution hazard models, and Gray’s tests were performed using the “tidycmprsk” package^39^ in R.

## Data availability

The data analyzed in this study are not publicly available because of ethical restrictions. However, they may be made available upon request after approval by

the Ethics Committee of Iwate Medical University, the Ethics Committee of Tohoku University, and the Materials and Information Distribution Review Committee of the TMM Project.

## Results

### Polygenic score and baseline characteristics

The baseline characteristics of the study participants stratified by PGS quintiles are presented in Table 1. A significant inverse trend was observed between PGS quintiles and age (*P_trend_* =0.020). This association remained robust after adjusting for the first five PCs (Table S1) (*P_trend_* =0.007). The lower mean age and lower proportion of males observed in the higher PGS quintiles may differences in participant characteristics across genetic risk groups.

**Table 1.**
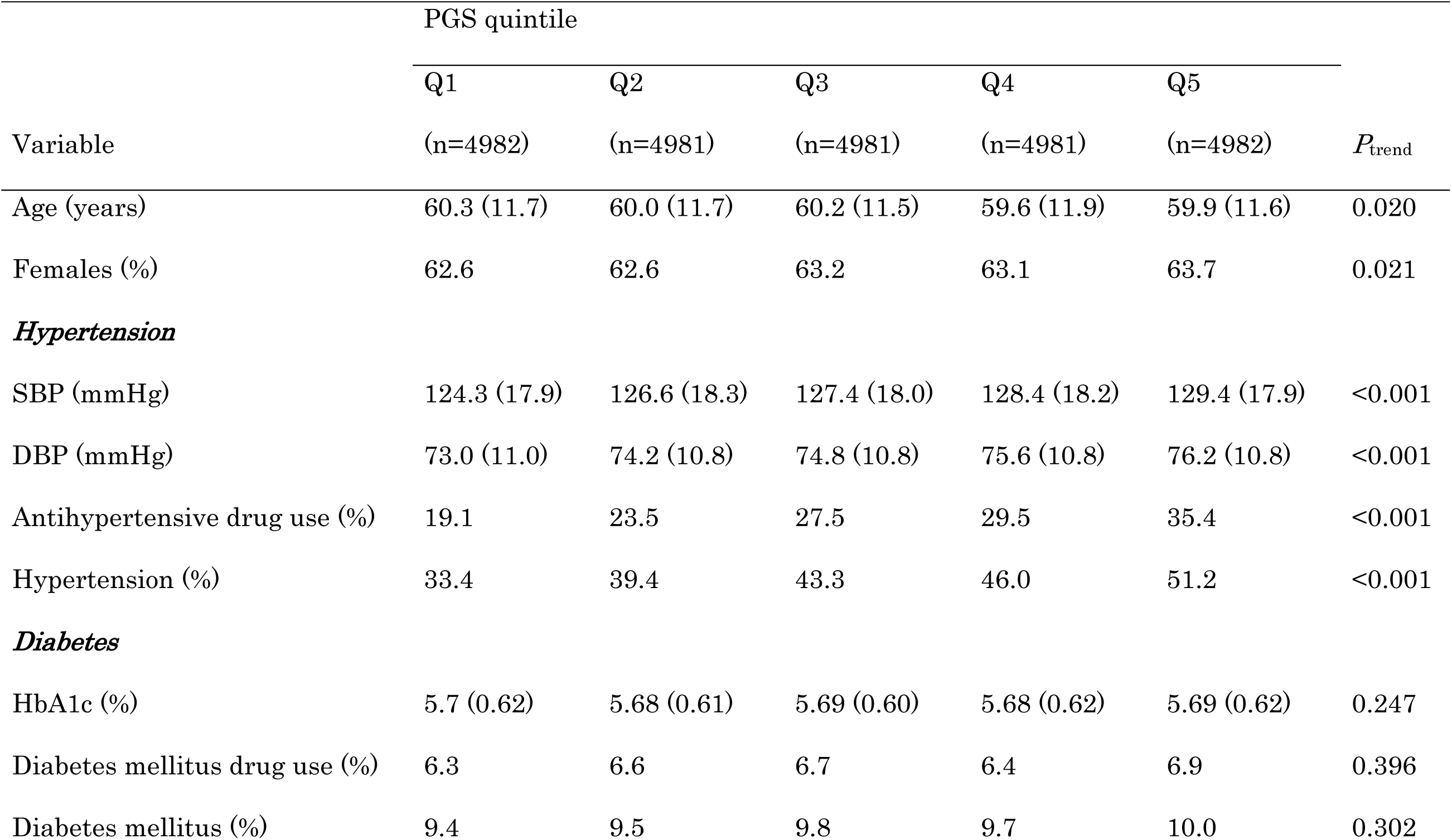
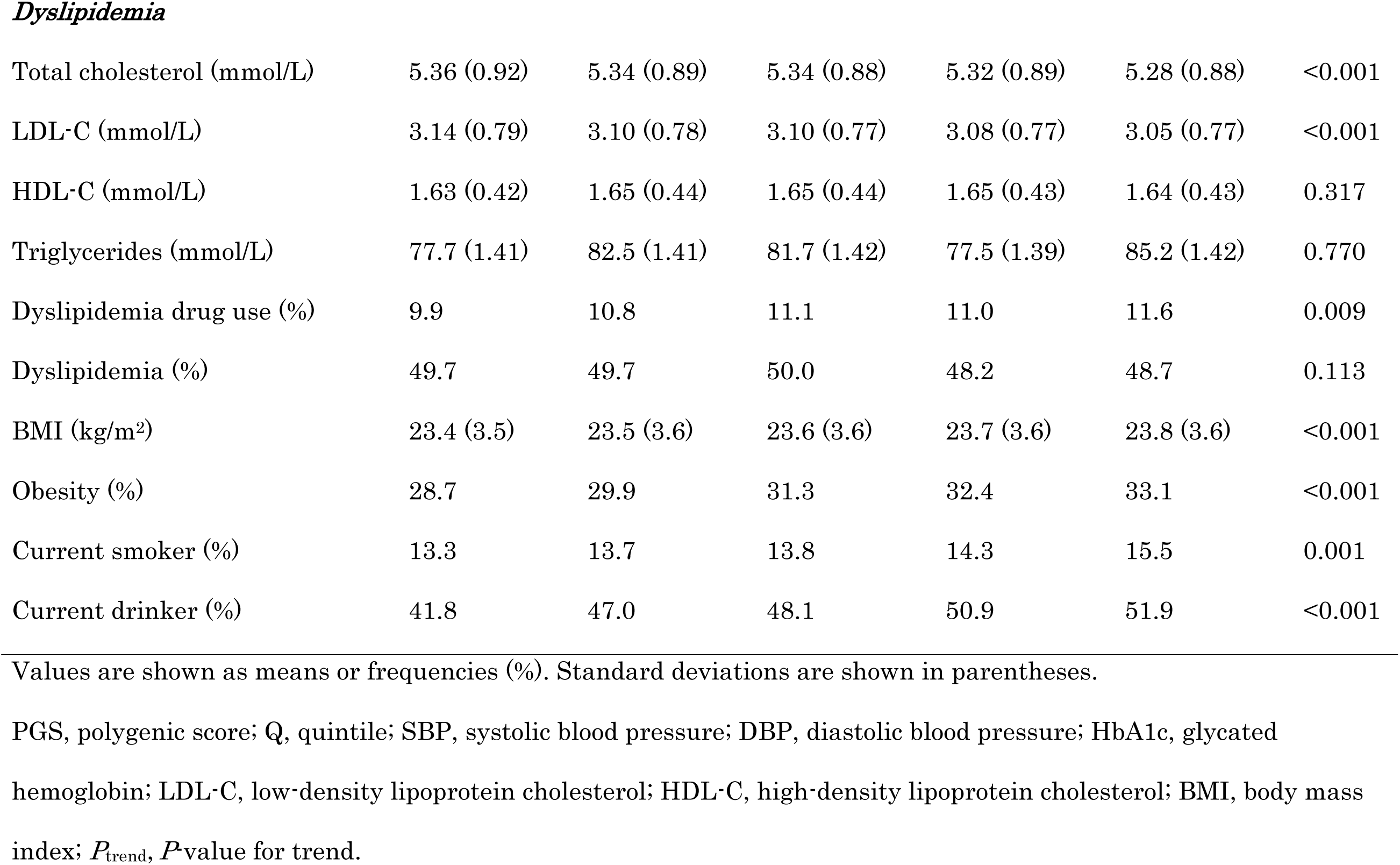
Baseline characteristics of study participants according to PGS quintiles.

| | PGS quintile | | | | | $P_{\text{trend}}$ |
| --- | --- | --- | --- | --- | --- | --- |
|  | Q1 | Q2 | Q3 | Q4 | Q5 |  |
| Variable | (n=4982) | (n=4981) | (n=4981) | (n=4981) | (n=4982) |  |
| Age (years) | 60.3 (11.7) | 60.0 (11.7) | 60.2 (11.5) | 59.6 (11.9) | 59.9 (11.6) | 0.020 |
| Females (%) | 62.6 | 62.6 | 63.2 | 63.1 | 63.7 | 0.021 |
| ***Hypertension*** |  |  |  |  |  |  |
| SBP (mmHg) | 124.3 (17.9) | 126.6 (18.3) | 127.4 (18.0) | 128.4 (18.2) | 129.4 (17.9) | <0.001 |
| DBP (mmHg) | 73.0 (11.0) | 74.2 (10.8) | 74.8 (10.8) | 75.6 (10.8) | 76.2 (10.8) | <0.001 |
| Antihypertensive drug use (%) | 19.1 | 23.5 | 27.5 | 29.5 | 35.4 | <0.001 |
| Hypertension (%) | 33.4 | 39.4 | 43.3 | 46.0 | 51.2 | <0.001 |
| ***Diabetes*** |  |  |  |  |  |  |
| HbA1c (%) | 5.7 (0.62) | 5.68 (0.61) | 5.69 (0.60) | 5.68 (0.62) | 5.69 (0.62) | 0.247 |
| Diabetes mellitus drug use (%) | 6.3 | 6.6 | 6.7 | 6.4 | 6.9 | 0.396 |
| Diabetes mellitus (%) | 9.4 | 9.5 | 9.8 | 9.7 | 10.0 | 0.302 |
| *Dyslipidemia* |  |  |  |  |  |  |
| Total cholesterol (mmol/L) | 5.36 (0.92) | 5.34 (0.89) | 5.34 (0.88) | 5.32 (0.89) | 5.28 (0.88) | <0.001 |
| LDL-C (mmol/L) | 3.14 (0.79) | 3.10 (0.78) | 3.10 (0.77) | 3.08 (0.77) | 3.05 (0.77) | <0.001 |
| HDL-C (mmol/L) | 1.63 (0.42) | 1.65 (0.44) | 1.65 (0.44) | 1.65 (0.43) | 1.64 (0.43) | 0.317 |
| Triglycerides (mmol/L) | 77.7 (1.41) | 82.5 (1.41) | 81.7 (1.42) | 77.5 (1.39) | 85.2 (1.42) | 0.770 |
| Dyslipidemia drug use (%) | 9.9 | 10.8 | 11.1 | 11.0 | 11.6 | 0.009 |
| Dyslipidemia (%) | 49.7 | 49.7 | 50.0 | 48.2 | 48.7 | 0.113 |
| BMI (kg/m<sup>2</sup>) | 23.4 (3.5) | 23.5 (3.6) | 23.6 (3.6) | 23.7 (3.6) | 23.8 (3.6) | <0.001 |
| Obesity (%) | 28.7 | 29.9 | 31.3 | 32.4 | 33.1 | <0.001 |
| Current smoker (%) | 13.3 | 13.7 | 13.8 | 14.3 | 15.5 | 0.001 |
| Current drinker (%) | 41.8 | 47.0 | 48.1 | 50.9 | 51.9 | <0.001 |

Values are shown as means or frequencies (%). Standard deviations are shown in parentheses.
PGS, polygenic score; Q, quintile; SBP, systolic blood pressure; DBP, diastolic blood pressure; HbA1c, glycated hemoglobin; LDL-C, low-density lipoprotein cholesterol; HDL-C, high-density lipoprotein cholesterol; BMI, body mass index; $P_{\text{trend}}$ , $P$ -value for trend.

Among the physiological risk factors for stroke, SBP and DBP showed significant positive trends with increasing PGS quintile, whereas TC and LDL-C levels showed significant inverse trends (Table 1). Lifestyle-related factors, including obesity, current smoking, and current alcohol consumption, were also significantly associated with higher PGS levels. These associations remained significant after adjusting for genetic PCs, age, and sex (Table S1), suggesting that the relationships between these modifiable risk factors and genetic susceptibility to stroke were largely independent of these potential confounding factors.

### Risk of incident stroke according to genetic predisposition

During follow-up, 246 stroke events occurred (Table 2). The incidence of stroke increased progressively across PGS quintiles, ranging from 1.64 per 1,000 person-years in the lowest quintile (Q1) to 2.79 per 1,000 person-years in the highest quintile (Q5).

**Table 2.** Risk of incident stroke according to PGS quintiles

| PGS<br>quintile | Stroke<br>events | PYs at<br>risk | Incidence rate<br>(per 10<sup>3</sup> PYs) | HR (95% CI)<sup>a</sup> | *P*-value | *P*<sub>trend</sub><sup>a</sup> |
| --- | --- | --- | --- | --- | --- | --- |
| Q1 | 40 | 24329.6 | 1.64 | 0.93 (0.61–1.42) | 0.734 |  |
| Q2 | 44 | 24390.9 | 1.80 | 1.00 (0.66–1.53) | 0.968 |  |
| Q3 | 44 | 24410.7 | 1.80 | 1.00 (Ref.) | Ref. | 0.003 |
| Q4 | 50 | 24354.5 | 2.05 | 1.19 (0.80–1.79) | 0.391 |  |
| Q5 | 68 | 24357.6 | 2.79 | 1.60 (1.10–2.34) | 0.015 |  |

<sup>a</sup> Estimated by the Cox proportional hazards model adjusted by age and sex.
PGS, polygenic score; Q, quintile; CI, confidence interval; HR, hazard ratio; PYs, person-years; Ref, reference; *P*<sub>trend</sub>, *P*-value for trend.

After adjustment for age and sex, participants in the highest PGS quintile (Q5) had a significantly higher risk of incident stroke than those in the middle quintile (Q3) (HR, 1.60; 95% CI, 1.10–2.34). This association remained robust after further adjustment for the first five genetic PCs (Table S2).

### Genetic risk and modifiable risk factors

Multiple modifiable risk factors were significantly associated with an increased risk of incident stroke, including hypertension (HR, 1.89; 95% CI, 1.44–2.50), diabetes mellitus (HR, 1.44; 95% CI, 1.03–1.99), dyslipidemia (HR, 1.48; 95% CI, 1.14–1.91), obesity (HR, 1.38; 95% CI, 1.07–1.78), and current smoking (HR, 1.73; 95% CI, 1.24–2.41). Notably, the magnitude of stroke risk associated with these modifiable risk factors was comparable to the excess risk observed between participants in the intermediate (Q3) and high (Q5) genetic risk groups (HR, 1.60; 95% CI, 1.10–2.34) (Figure 1; Table S3).

**Figure 1.**
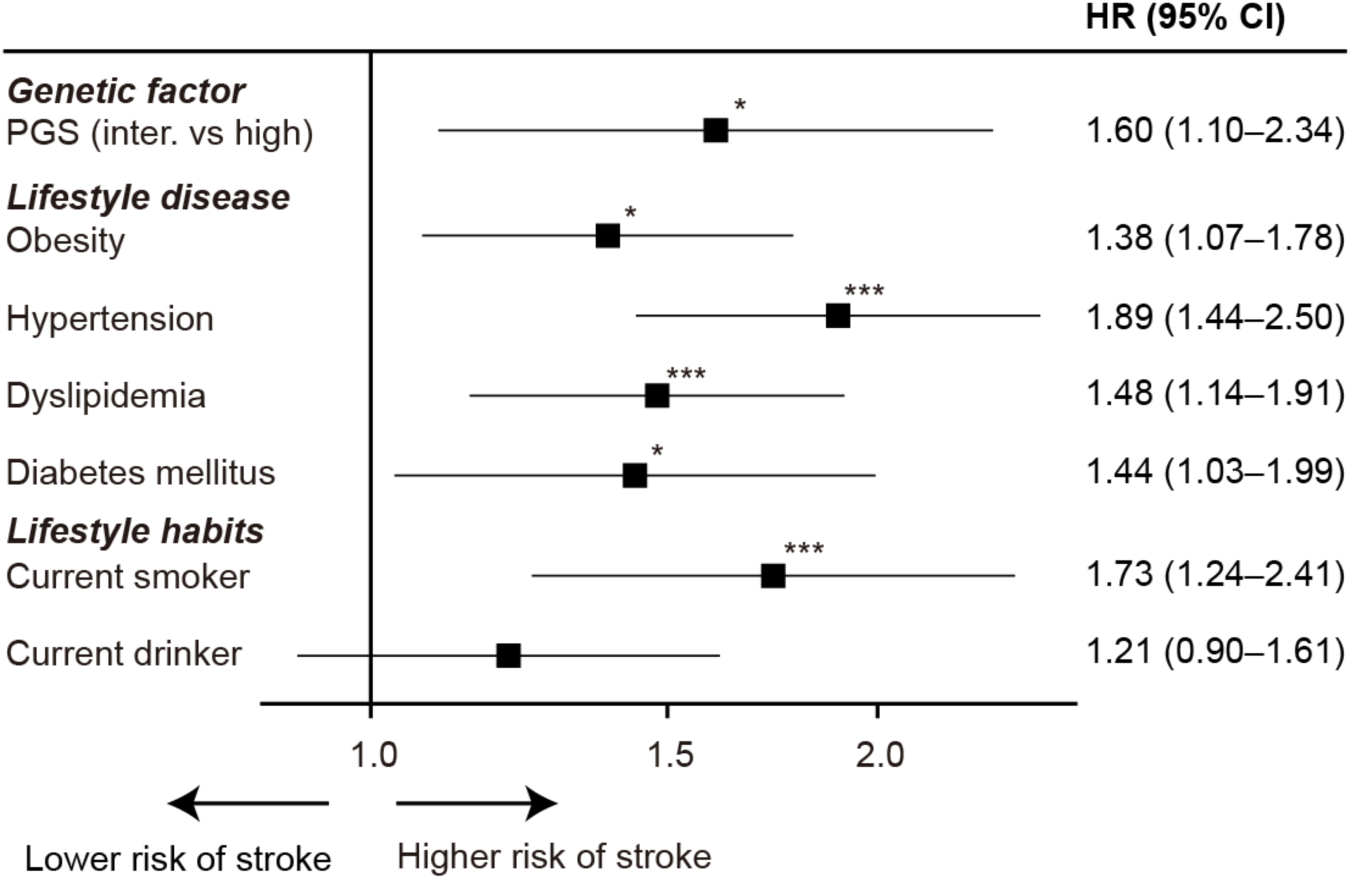
Risk of incident stroke according to genetic risk and modifiable risk factors. The logarithmic x-axis indicates age- and sex-adjusted HRs with 95% CIs. For the PGS, HRs were calculated for participants in the highest quintile (Q5), with those in the middle quintile (Q3) serving as the reference group. Detailed information on the number of participants, number of events, and follow-up duration is presented in Table S3. PGS, polygenic score; HR, hazard ratio; CI, confidence interval; Q, quintile. *P<0.05; \*\**P*<0.01; \*\*\**P*<0.005.

Among participants in the intermediate genetic risk group (Q3), the risk of incident stroke was significantly increased in the presence of hypertension (HR, 2.17; 95% CI, 1.11–4.25) and diabetes mellitus (HR, 2.10; 95% CI, 1.04–4.27) (Figure 2; Table S4). In contrast, participants in the high genetic risk group (Q5) exhibited a significantly higher risk of stroke than individuals with intermediate genetic risk (Q3) who did not have the corresponding modifiable risk factor.

**Figure 2.**
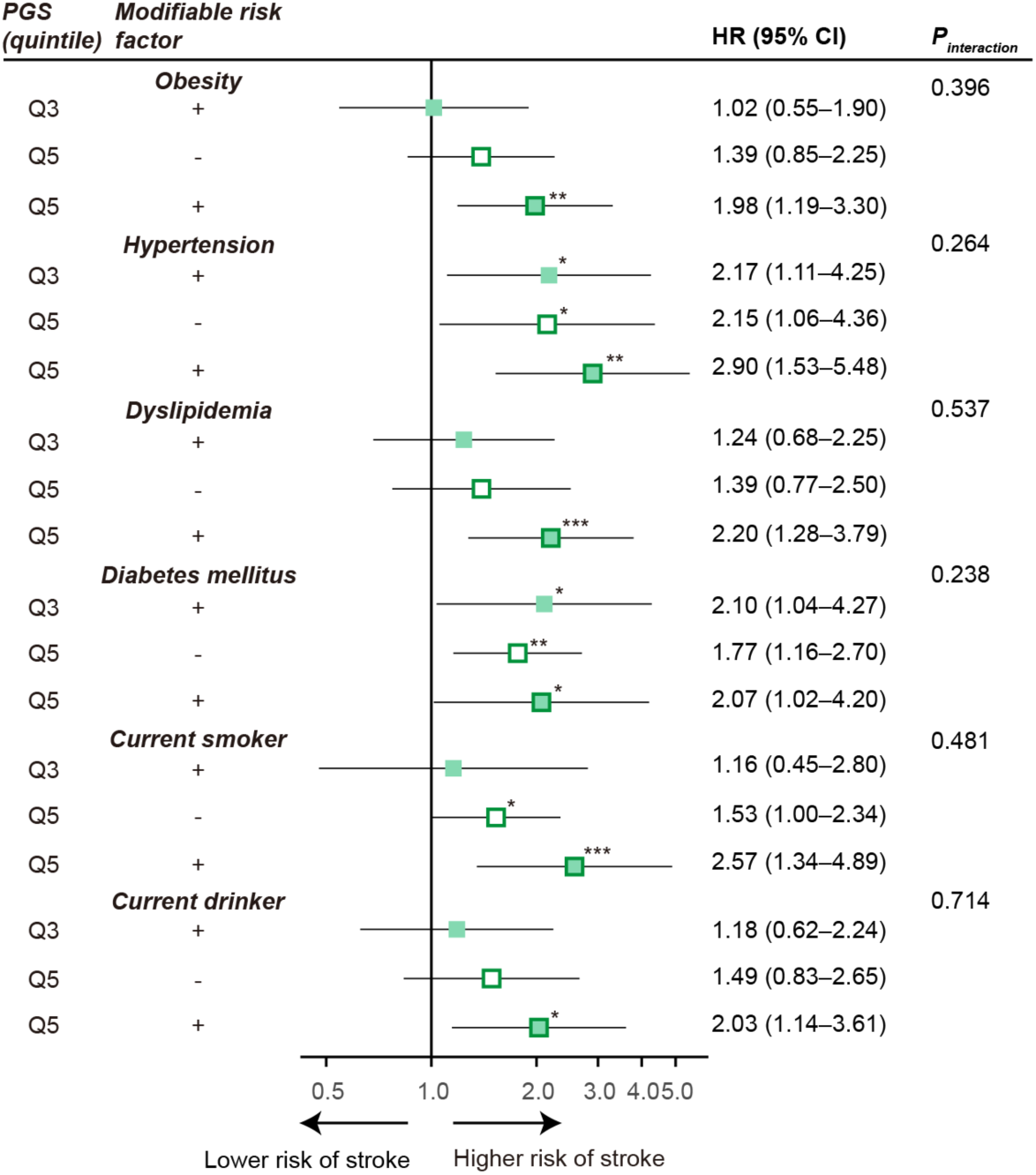
Risk of incident stroke according to combined genetic and modifiable risk factors. The logarithmic x-axis indicates age- and sex-adjusted HRs with 95% CIs. HRs for each modifiable risk factor were estimated using participants without the corresponding modifiable risk factor and with intermediate genetic risk (Q3) as the reference group. Plus (+; filled boxes) and minus (−; open boxes) symbols indicate the presence and absence of the corresponding modifiable risk factor, respectively. Results for the high genetic risk group (Q5) are indicated by boxes with green outlines. Interactions between the PGS and modifiable risk factors were assessed using analysis of variance (ANOVA) (*P* for interaction). Detailed information is presented in Table S4. PGS, polygenic score; HR, hazard ratio; CI, confidence interval; Q, quintile. \**P*<0.05; \*\**P*<0.01; \*\*\**P*<0.005.

Notably, within the high genetic risk group, stroke risk was further increased in participants with dyslipidemia (HR, 2.20; 95% CI, 1.28–3.79), obesity (HR, 1.98; 95% CI, 1.19–3.30), current smoking (HR, 2.57; 95% CI, 1.34–4.89), and current alcohol consumption (HR, 2.03; 95% CI, 1.14–3.61).

The number of modifiable risk factors was significantly associated with incident stroke risk in the high genetic risk group (HR per additional risk factor (trend), 1.35; 95% CI, 1.11–1.64; *P_trend_*=0.003) but not in the intermediate genetic risk group (HR per additional risk factor (trend), 1.27; 95% CI, 0.98–1.64; *P_trend_*=0.075) (Figure 3; Table S5). Similarly, after stratification according to both PGS level and the number of modifiable risk factors (0–1, 2–3, and ≥4 categories, defined to achieve comparable sample sizes), participants with high genetic risk and ≥4 modifiable risk factors exhibited a significantly increased risk of stroke (HR, 2.76; 95% CI, 1.28–5.92; *P*=0.009). No statistically significant interaction was observed between PGS and the number of modifiable risk factors (*P_interaction_*>0.05). Comparable results were obtained using the Fine–Gray subdistribution hazard model, accounting for all-cause mortality as a competing risk factor for stroke (Table S6). Participants with high genetic risk and ≥4 modifiable risk factors remained at significantly higher risk of stroke than the reference group (HR, 2.78; 95% CI, 1.26–6.13; *P*=0.011).

**Figure 3.**
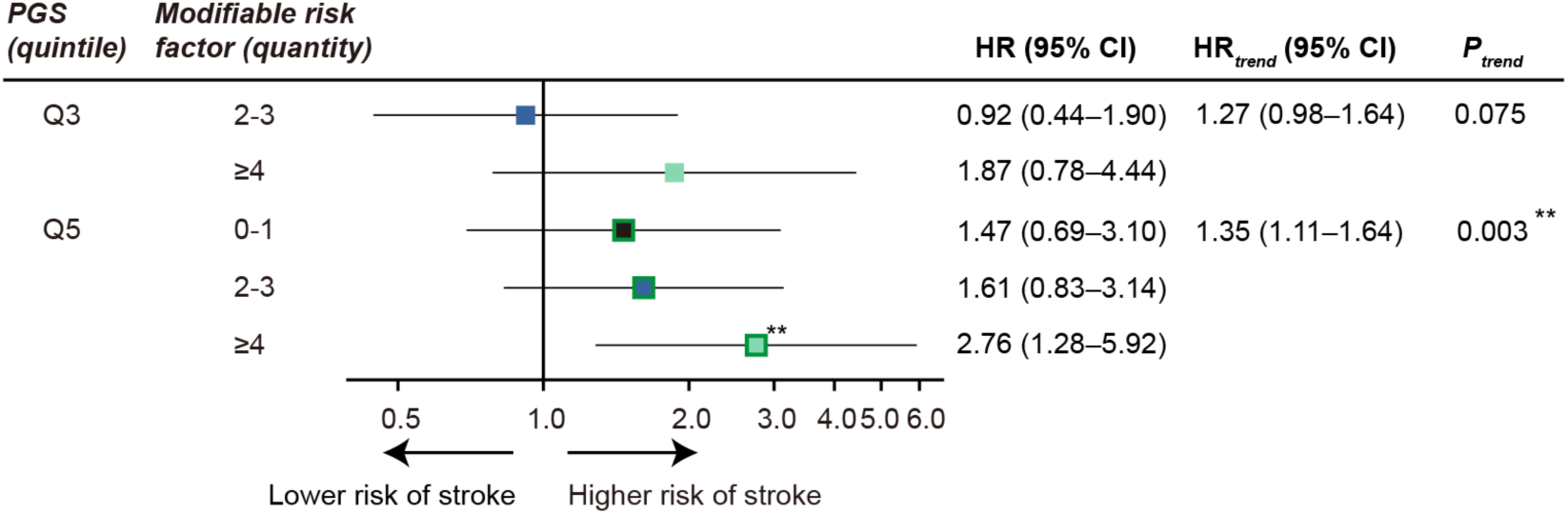
Risk of incident stroke according to genetic risk and the number of modifiable risk factors. HRs according to the number of modifiable risk factors (0–1, 2–3, and ≥4) were estimated using participants with intermediate genetic risk (Q3) and 0–1 modifiable risk factor as the reference group. The categories of modifiable risk factors are represented by black-, blue-, and light green-filled boxes, respectively. Results for the high genetic risk group (Q5) are indicated by boxes with green outlines. See the legend of Figure 2 for additional details. Detailed information on the number of participants, number of events, and follow-up duration is presented in Table S5. PGS, polygenic score; HR, hazard ratio; CI, confidence interval; Q, quintile. \**P*<0.05; \*\**P*<0.01; \*\*\**P*<0.005.

The combination of PGS category and the number of modifiable risk factors significantly influenced the cumulative incidence of stroke (*P* for Gray’s test [*P_Gray_*] <0.001), based on analyses accounting for all-cause mortality before stroke onset as a competing risk (Figure 4; Table S7). Under this competing-risk framework, participants with high genetic risk and ≥4 modifiable risk factors had estimated cumulative stroke incidences of 1.02% (95% CI, 0.39–2.23) and 22.6% (95% CI, 5.4–46.9) at 70 and 80 years of age, respectively (Table S7). In contrast, the corresponding incidences among participants with high genetic risk and 0‒1 modifiable risk factors were 0.6% (95% CI, 0.25–1.29) and 6.18% (95% CI, 3.02– 10.9) at 70 and 80 years of age, respectively.

**Figure 4.**
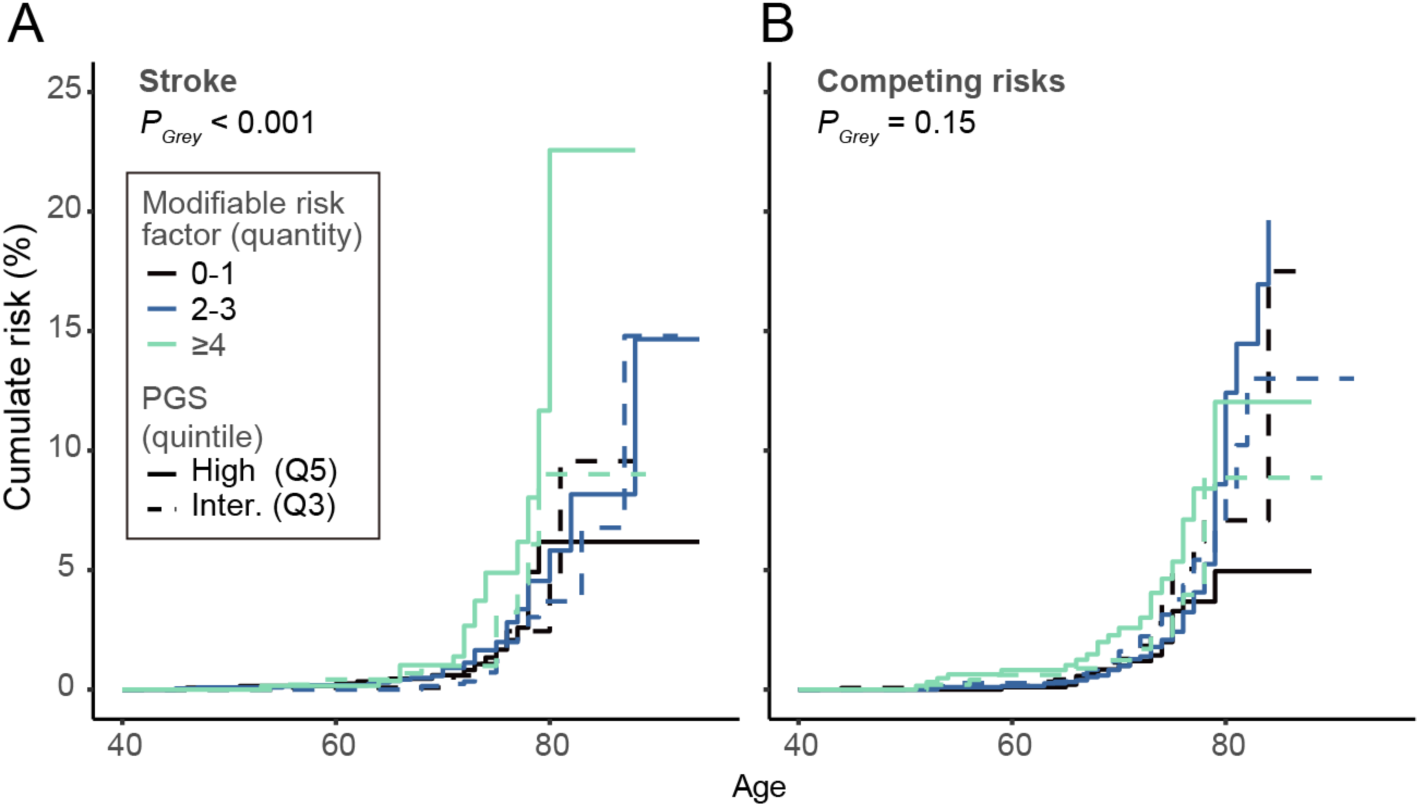
Cumulative incidence of stroke accounting for all-cause mortality as a competing risk. Cumulative incidence of stroke (A) was estimated using the Fine–Gray model, accounting for all-cause mortality as a competing risk (B). The x-axis indicates age, and the y-axis indicates the estimated cumulative incidence of stroke, accounting for all-cause mortality as a competing risk. Colors represent the number of modifiable risk factors. Solid and dotted lines indicate high (Q5) and intermediate PGS (Q3; reference) groups, respectively. Detailed information, including the standard errors of the estimates and numbers at risk and events, is presented in Table S7. PGS, polygenic score; Q, quintile.

Similar trends were observed in the female subgroup (*P_Gray_* for stroke=0.017; *P_Gray_* for competing risk=0.89; see Table S8 for age-specific cumulative incidence estimates), whereas this association was not statistically significant in the male subgroup (*P_Gray_* for stroke=0.18; *P_Gray_* for competing risk=0.30; see Table S9).

## Discussion

Stroke predominantly occurs later in life, indicating that individuals carry their genetic susceptibility for several decades before disease onset. Therefore, prevention strategies based on genetic risk require long-term management of modifiable risk factors throughout life while balancing preventive benefits and individual burden or cost. Additional evidence is required to establish practical and cost-effective prevention strategies tailored to individual genetic risk profiles.

The findings of the present study suggest that the relative importance of modifiable risk factors for primary prevention may differ according to an individual’s genetic predisposition. While the management of hypertension and diabetes mellitus may be particularly important among participants with intermediate genetic risk, those with high genetic risk may require a broader approach to risk factor management, including the control of dyslipidemia and obesity, as well as smoking and alcohol cessation.

Furthermore, our findings are broadly consistent with those reported by Hachiya et al.^17^, who suggested that reducing the number of modifiable risk factors may attenuate the overall stroke risk, even among individuals with high genetic susceptibility. Moreover, analyses using cumulative incidence functions and Fine–Gray subdistribution hazard models that account for competing risks yielded similar results, supporting the robustness of our findings.

The present findings suggest that the multifactorial threshold model may provide a practical framework for understanding the interplay between genetic predisposition and environmental risk factors in stroke prevention^40^. In this context, PGS may enhance the efficiency of conventional public health strategies by identifying individuals at high genetic risk who may benefit from intensive preventive interventions and personalized lifestyle guidance as part of risk communication^41^.

The GIGASTROKE PGS model used in the present study integrates genetic susceptibility across multiple stroke-related traits and risk factors, including atrial fibrillation, coronary artery disease, diabetes mellitus, SBP, DBP, TC, LDL-C, HDL-C, TG, BMI, height, and smoking behavior. Therefore, among individuals with a high PGS, avoiding smoking may help effectively mitigate one of the major pathways linking genetic susceptibility to stroke risk. Future interventional studies evaluating lifestyle modifications based on genetic risk are warranted.

This study has several limitations. First, the number of incident stroke events was relatively limited for a genetic epidemiological study, which may have reduced the statistical power, particularly for cumulative incidence analyses stratified by age, PGS category, and the number of modifiable risk factors. Second, age and sex distributions differed across the PGS quintiles, suggesting potential bias. This may partly reflect the exclusion of individuals with a history of stroke at baseline, as well as the recruitment framework, which was based primarily on Japan’s National Health Insurance system. Because this framework excludes individuals covered exclusively by employee health insurance, it may have introduced a bias toward females and retired individuals. Consequently, older males with high genetic risk may have been underrepresented in the cohort, potentially resulting in an underestimation of stroke incidence in the higher PGS groups, particularly in cumulative incidence analyses.

In conclusion, the accumulation of modifiable risk factors substantially influenced stroke risk even among individuals with high genetic susceptibility. Differences in the magnitude of risk associated with individual modifiable factors may help inform the prioritization of preventive strategies according to genetic risk profiles and contribute to the development of personalized approaches to stroke prevention.

## Data Availability

The data analyzed in this study are not publicly available because of ethical restrictions. However, they may be made available upon request after approval by the Ethics Committee of Iwate Medical University, the Ethics Committee of Tohoku University, and the Materials and Information Distribution Review Committee of the TMM Project.

## Acknowledgments

We thank Dr. Masato Nagai for his contribution to the curation and management of follow-up data. During the preparation of this manuscript, the authors used ChatGPT (OpenAI) for English language editing. The authors reviewed and edited all the generated content and take full responsibility for the final manuscript.

## Sources of Funding

This study was supported by the TMM Project (Special Account for the Reconstruction of the Great East Japan Earthquake) of the Ministry of Education, Culture, Sports, Science and Technology (MEXT) and the Japan Agency for Medical Research and Development (AMED) under grant number JP23tm0124006. The TMM Project supercomputer system, under grant number JP21tm0424601, was used for the data analyses. YS and AS were supported by the Japan Society for the Promotion of Science (JSPS) KAKENHI grants 23K09747, 26K13245 and 26K13245.

## Declaration of interests

T.H. receives personal fees from Genome Analytics Japan Inc. outside the submitted work and has a patent (PCT/JP2016/080192) pending and a patent (JP6312253B2) licensed. All other authors declare no conflicts of interest.

## Author contributions

Conceptualization: YS, TH, and AS Data curation: YOY, MN, SO, YK, HA, Y Kotozaki, SK, SM, HO, NN, MS, and KT. Formal analysis: YS and TH

Writing – original draft: YS and TH Writing, review, and editing: YS, TH, YOY, MN, SO, Y K, HA, YKotozaki, SK, SM, HO, NN, MS, KT, and AS Supervision: YS, TH, and AS

## Non-standard Abbreviations and Acronyms

PGS, polygenic score; Q, quintile; TMM, Tohoku Medical Megabank; TMM CommCohort, Tohoku Medical Megabank Project community-based cohort

## Notes

### Author Declarations

This study was approved by the Institutional Review Board of Iwate Medical University (Approval Nos. HG H25-2, HG2018-004, HG2021-011, and 2014-0001). Written informed consent was obtained from all the participants. All study procedures were conducted in accordance with the principles of the Declaration of Helsinki.

